# Examining impacts of approval of home use of misoprostol in England on access to medical abortion

**DOI:** 10.1101/2022.03.28.22273043

**Authors:** Maria Lewandowska, Daniel J Carter, Patricia A. Lohr, Kaye Wellings

## Abstract

**Objectives:** To assess the impact of the December 2018 approval of home administration of misoprostol in England on access to medical abortion.

**Design:** Time series analysis

**Setting:** British Pregnancy Advisory Service (BPAS), independent-sector abortion provider in England

**Participants:** 145,529 abortions carried out by BPAS across England between 2018 and 2019.

**Intervention:** Approval of home administration of misoprostol in early medical abortions (EMA) in December 2018

**Main outcome measure:** Gestational age at abortion and EMAs as a proportion of all abortions. The analysis was stratified by key sociodemographic characteristics to assess differential impacts of the approval

**Results:** 99,008 abortions took place in the period before the approval or during its implementation phase (January 2018 – June 2019) and 46,521 took place after (July 2019 – Dec 2019). Compared to if former trends had continued, the actual proportion of EMAs was 4.2% higher in December 2019 and the mean gestational age 3.4 days lower.

**Conclusion:** Following the approval of home use of misoprostol, we saw an acceleration in the trends towards increasing proportion of all abortions that were EMAs and decreasing gestational age at abortion, especially in more deprived areas of England. Some inequities remain across race/ethnicity groups that require further investigation. Policymakers should take the positive results of this study into consideration when reviewing rules for home management of medical abortions, including with home use of mifepristone.

**What is already known on this topic:** In 2018 in England, a woman’s “home” was designated as a class of place where misoprostol could be used to induce abortion up to 10 weeks’ gestation following administration of mifepristone in a medical facility. This model of abortion care has been shown in numerous international studies to be highly effective, safe, and preferred by women over in-clinic use. Existing data anticipated positive clinical and acceptability outcomes with implementation of home use, but whether or how the change would impact access particularly in relation to barriers such as area-level deprivation, race/ethnicity, and disability was uncertain.

**What this study adds:** The approval of home use of misoprostol as part of a medical abortion regimen in England was associated with material and equitable improvements in abortion access as evidenced by a higher proportion of medical abortions provided, lower gestational age at treatment, and higher odds of having a medical abortion across all racial/ethnic groups and socioeconomic groups. Pre-approval trends toward greater uptake of medical abortion and declining gestational age were accelerated post-approval and were greatest in the most deprived quintiles but not across all racial/ethnic groups.

**Patient and Public Involvement Statement:** This study was a quantitative data analysis of existing clinical data and patients were not directly involved in the research.

**Authors’ note on terminology:** The authors would like to note that abortions are experienced not only by cis-women, but also by trans, non-binary and intersex people, who should be recognised and treated as equal recipients of abortion care. The term ‘women’ will be used in this project for simplicity and in acknowledgment of the fact that the majority of the patients identify as women.

## Background

Medical abortion involves the administration of mifepristone, followed 24-48 hours later by misoprostol, resulting in loss of the pregnancy in a process similar to miscarriage. (1) The proportion of medical abortions in Britain has been increasing since the introduction of mifepristone in the early 1990s. In 2001, medical abortion accounted for only 12% of all abortions across England and Wales. (2,3) By 2011, this proportion had risen to 47%, and by 2019, to 73%. (4,5)

Early medical abortion (EMA) refers to the use of mifepristone and misoprostol in the first 10-12 weeks of pregnancy. In many parts of the world, self-administration of misoprostol at home has been the standard model of care, in line with evidence demonstrating it to be safe, effective, and acceptable. (6,7) Until 2017, those seeking EMA in Britain were legally required to attend an approved clinic or NHS hospital to have these medications administered. (8,9) In almost all cases, the patients would not remain in the clinical setting after receiving the medications but would return home to complete the abortion.

Home administration of medical abortion has been repeatedly proposed in the early stages of pregnancy as a means of improving access to care and mitigating psychological, financial, and logistical burdens. It has several advantages: it allows for increased privacy, affords opportunities for support from family and friends and provides greater comfort. (10–13) The alternative -travelling home after administration of misoprostol in a clinical setting - increases travel time and associated expenses, can incur income loss, and carries the risk of causing distress in the event of onset of bleeding and pain during the journey. (3,10,13–16)

Increasing access to medical abortion can also result in abortions being provided earlier in pregnancy by mechanisms including both increased provider capacity and improved patient experience. While abortion is a very safe procedure overall, the risk of potential complications increases with every subsequent week of gestation. (1) Earlier abortions minimise the risk of adverse events and improve women’s experience of abortion care. There is also strong evidence that earlier abortions are more cost-effective for health systems: the savings a consequence of choosing medical over surgical abortions and preventing complications. (1,17,18)

In December 2018, the Secretary of State for Health in England approved “home” as a place where misoprostol as part of medical abortion regimen could lawfully be administered. This measure, conforming with the World Health Organization (WHO) guidelines, (19) brought England in line with Wales, where home use had been approved in June 2018, and with Scotland, where it was approved in October 2017. (20) The approval of home use in England and Wales is limited to early medical abortions (EMA), defined as under 10 weeks’ gestation. In 2019, 36% of medical abortions in England and Wales were carried out with misoprostol administered at home. (5)

In this study, we examined the impacts of the December 2018 ruling permitting home administration of misoprostol. We analysed routinely collected data on abortion to explore changes in the proportion of all abortions carried out by EMA, and in the gestational age at which abortions were carried out, before and after the ruling. We also examined whether any such changes varied by key population characteristics related to inequality, to understand effects by area-level deprivation, race/ethnicity, and disability.

## Methods

### Dataset

Three-quarters of abortions in England and Wales are provided by independent sector clinics working under NHS contracts. (5) We used data from one of these, the British Pregnancy Advisory Service (BPAS), which provides almost 33% of all abortions in Britain. (21) We extracted anonymised aggregate data from BPAS’ Booking and Invoicing System (BIS) on all abortions provided between January 2015 and December 2019. The proportion of EMAs at BPAS has been increasing logarithmically since 2015 (see Figure S1). We therefore restricted our analysis to the time when the increase in this proportion was linear and stationary: from January 2018 to December 2019.

### Research question

We asked whether the approval permitting home administration of misoprostol was associated with a change in the ratio of EMAs to late medical or surgical abortions, in gestational age at treatment, and whether any observed changes varied with patient characteristics.

### Dataset

We used the restricted sample from January 2018 to December 2019 and described it in terms of abortion method, gestational age, past experience of abortion and distribution of demographic characteristics. In this paper, we use the term EMA as under 10 weeks’ gestation. Since whether the impacts of the approval on access to care vary in line with social inequities is of considerable public health interest, we conducted three further stratified analyses to examine differential effects by area-level deprivation, race/ethnicity, and disability. Deprivation was derived from the first three letters of patients’ postcode linked to the Index of Multiple Deprivation in England (IMD),(22) and divided into quintiles. Race/ethnicity was determined by self-report based on the NHS ethnic category code, (23) and disability was defined in accordance with the Equality Act 2010, as physical or mental impairment with a substantial, long-term negative effect on health. (24)

### Statistical analysis

We conducted a Mantel-Haenszel analysis to compare the prevalence of EMA in the period before and after the approval, stratified by patient subgroups, and reported proportions and crude odds ratios (ORs). We also conducted a univariable linear regression to provide mean gestational age at abortion, stratified by patient characteristics, and the risk difference pre-post the approval.

We then conducted an interrupted time series analysis (ITS) using segmented linear regression analyses. We examined the slope of fitted regression lines that represent the expected increase or decrease in gestational age or proportion of EMAs for each additional month in time. We specifically estimated the expected change in these slopes after the implementation of the new care model, anticipating an effect of the approval on the rate of change in our outcomes of interest. Given full implementation of the exposure was not achieved until approximately 6 months after the approval (see Figure S2), we include a 6-month transition period post-legislation as part of the pre-implementation period. Effect modification by social categories (deprivation, race/ethnicity, disability) was tested by introducing a three-way interaction term into the model to test the null hypothesis that the slope change post-approval was the same in each group. A Durbin-Watson test was conducted, and visual plots of autocorrelation functions were produced to examine whether the ITS assumption that each observation is not dependent on previous observations holds.

Change in slopes expected across the time period were plotted graphically. We also estimated average differences in outcome at the endpoint of December 2019 by comparing the observed outcomes to a counterfactual in the absence of intervention, that is, assuming that there was no slope change. All analyses presented in this paper were conducted in R v4.1.2 and the code is available post-publication at http://github.com/danieljcarter/bpas.

## Results

### Study population characteristics

Between January 2018 and December 2019, 145,548 abortions were conducted by BPAS, of which 102,592 (70.5%) were EMAs. The gestational age at abortion ranged from 21 days (3 weeks) to 168 days (24 weeks), with a median of 52 days (7 weeks and 3 days), and a mean of 59 days, rounded to the nearest whole number. The characteristics of the sample are provided in Table 1.

**Table 1.** Abortion method, gestational age and client characteristics at BPAS between January 2018 and December 2019. (N=145,529)

| **Characteristic** |  | **n (%) or median (IQR)** |
| --- | --- | --- |
| **Abortion method:** | **Late medical / surgical abortion** | 42937 (29.5%) |
|  | **EMA** | 102592 (70.5%) |
| **Gestational age (days)** |  | 52 (IQR: 45-63) |
| **Age (years)** | **<18** | 5949 (4.1%) |
|  | **18-25** | 58928 (40.5%) |
|  | **26-35** | 60687 (41.7%) |
|  | **36-45** | 19735 (13.6%) |
|  | **>45** | 230 (0.2%) |
| **Place of birth** | **England** | 110140 (75.7%) |
|  | **European Union** | 11597 (8.0%) |
|  | **Northern Ireland** | 245 (0.2%) |
|  | **Outside of EU** | 21906 (15.1%) |
|  | **Scotland** | 758 (0.5%) |
|  | **Wales** | 783 (0.5%) |
|  | **Missing** | 100 (<0.1%) |
| **Disability** | **No/Prefer not to say** | 140597 (96.6%) |
|  | **Yes** | 4932 (3.4%) |
| **First language** | **English** | 140917 (96.8%) |
|  | **Other than English** | 4612 (3.2%) |
| **Race/Ethnicity** | **Asian or Asian British** | 12571 (8.6%) |
|  | **Black or Black British** | 9180 (6.3%) |
|  | **White** | 113153 (77.8%) |
|  | **Mixed** | 6502 (4.5%) |
|  | **Other/not stated** | 4123 (2.8%) |
| **Religion** | **None** | 96463 (66.3%) |
|  | **Christian** | 30573 (21.0%) |
|  | **Muslim** | 7507 (5.2%) |
|  | **Prefer not to say** | 5236 (3.6%) |
|  | **Other** | 5750 (4.0%) |
| **Previous abortions** | **No previous abortions** | 88815 (61.0%) |
|  | **Previous abortions** | 56689 (39%) |
|  | **Missing** | 25 (<0.1%) |
| **Index of Multiple Deprivation** | **Least Deprived** | 27610 (19.0%) |
|  | **Less Deprived** | 28123 (19.3%) |
|  | **Middle Quintile** | 29094 (20.0%) |
|  | **More Deprived** | 29320 (20.1%) |
|  | **Most Deprived** | 31382 (21.6%) |

### Changes before and after the approval of home misoprostol

Following the approval and implementation of home misoprostol, EMA as a proportion of all abortions carried out by BPAS increased from 69.8% to 72.0% [1.12 (1.09, 1.14)] (Table 2a). Notable increases were seen across clients of all ages except those under 18, among whom the proportion undergoing EMA remained the lowest, and increased the least [pre: 63.0%; post: 64.5%, OR=1.07 (0.95-1.20)]. Increases were seen across all quintiles of deprivation, and across all specified race/ethnicity and religious groups. The proportion undergoing an EMA was lower among patients reporting a disability and the increase post-ruling was modest [pre: 62.5%; post: 64.5%; OR=1.09 (0.96, 1.23)]. The greatest change in odds of having an EMA occurred in the group with previous experience of abortion [pre: 69.0%; post: 71.4%; OR=1.23 (1.08, 1.17)]. Across all strata, a decrease in the likelihood of EMA was seen only among patients born in Northern Ireland [pre: 79.4%; post: 70.0%, OR=0.61 (0.32, 1.17)].

**Table 2a.**
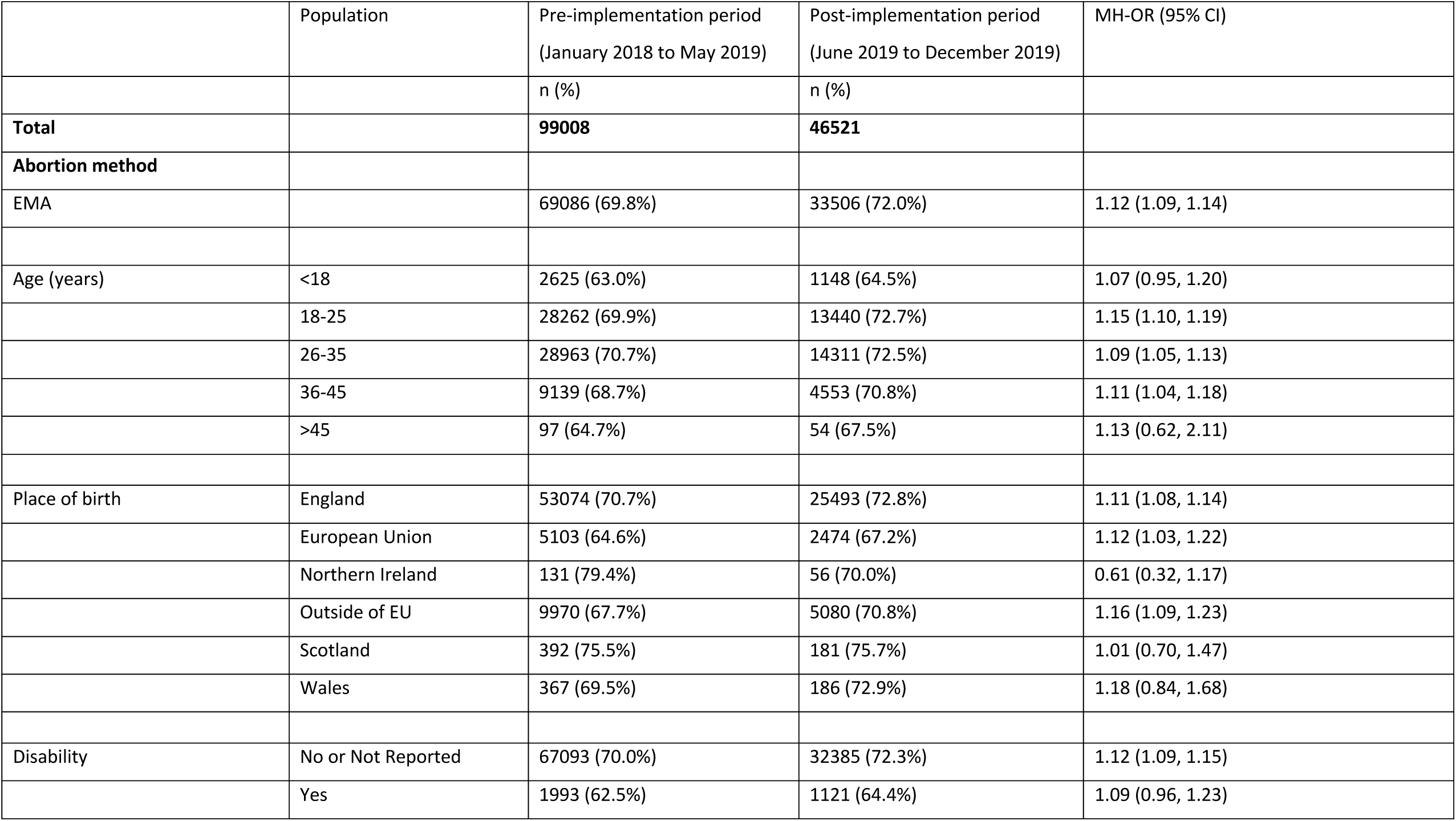
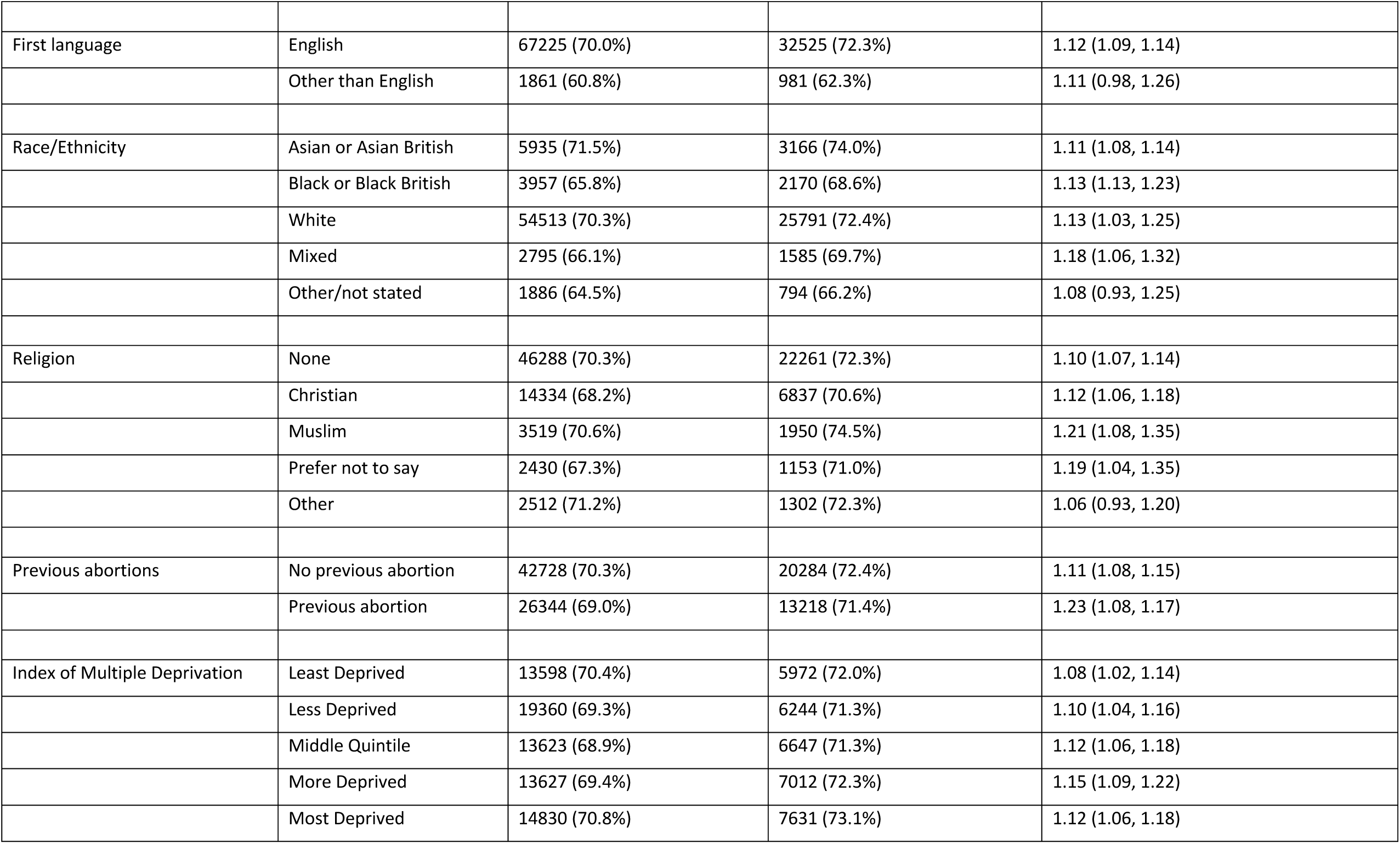
Proportion of patients having an EMA before and after full implementation of home use of misoprostol; stratum-specific odds ratios representing the change in odds of EMA pre and post. (N=145,529)

|  | Population | Pre-implementation period<br>(January 2018 to May 2019) | Post-implementation period<br>(June 2019 to December 2019) | MH-OR (95% CI) |
| --- | --- | --- | --- | --- |
|  |  | n (%) | n (%) |  |
| **Total** |  | **99008** | **46521** |  |
| **Abortion method** |  |  |  |  |
| EMA |  | 69086 (69.8%) | 33506 (72.0%) | 1.12 (1.09, 1.14) |
| **Age (years)** | <18 | 2625 (63.0%) | 1148 (64.5%) | 1.07 (0.95, 1.20) |
|  | 18-25 | 28262 (69.9%) | 13440 (72.7%) | 1.15 (1.10, 1.19) |
|  | 26-35 | 28963 (70.7%) | 14311 (72.5%) | 1.09 (1.05, 1.13) |
|  | 36-45 | 9139 (68.7%) | 4553 (70.8%) | 1.11 (1.04, 1.18) |
|  | >45 | 97 (64.7%) | 54 (67.5%) | 1.13 (0.62, 2.11) |
| **Place of birth** | England | 53074 (70.7%) | 25493 (72.8%) | 1.11 (1.08, 1.14) |
|  | European Union | 5103 (64.6%) | 2474 (67.2%) | 1.12 (1.03, 1.22) |
|  | Northern Ireland | 131 (79.4%) | 56 (70.0%) | 0.61 (0.32, 1.17) |
|  | Outside of EU | 9970 (67.7%) | 5080 (70.8%) | 1.16 (1.09, 1.23) |
|  | Scotland | 392 (75.5%) | 181 (75.7%) | 1.01 (0.70, 1.47) |
|  | Wales | 367 (69.5%) | 186 (72.9%) | 1.18 (0.84, 1.68) |
| **Disability** | No or Not Reported | 67093 (70.0%) | 32385 (72.3%) | 1.12 (1.09, 1.15) |
|  | Yes | 1993 (62.5%) | 1121 (64.4%) | 1.09 (0.96, 1.23) |
| First language | English | 67225 (70.0%) | 32525 (72.3%) | 1.12 (1.09, 1.14) |
|  | Other than English | 1861 (60.8%) | 981 (62.3%) | 1.11 (0.98, 1.26) |
| Race/Ethnicity | Asian or Asian British | 5935 (71.5%) | 3166 (74.0%) | 1.11 (1.08, 1.14) |
|  | Black or Black British | 3957 (65.8%) | 2170 (68.6%) | 1.13 (1.13, 1.23) |
|  | White | 54513 (70.3%) | 25791 (72.4%) | 1.13 (1.03, 1.25) |
|  | Mixed | 2795 (66.1%) | 1585 (69.7%) | 1.18 (1.06, 1.32) |
|  | Other/not stated | 1886 (64.5%) | 794 (66.2%) | 1.08 (0.93, 1.25) |
| Religion | None | 46288 (70.3%) | 22261 (72.3%) | 1.10 (1.07, 1.14) |
|  | Christian | 14334 (68.2%) | 6837 (70.6%) | 1.12 (1.06, 1.18) |
|  | Muslim | 3519 (70.6%) | 1950 (74.5%) | 1.21 (1.08, 1.35) |
|  | Prefer not to say | 2430 (67.3%) | 1153 (71.0%) | 1.19 (1.04, 1.35) |
|  | Other | 2512 (71.2%) | 1302 (72.3%) | 1.06 (0.93, 1.20) |
| Previous abortions | No previous abortion | 42728 (70.3%) | 20284 (72.4%) | 1.11 (1.08, 1.15) |
|  | Previous abortion | 26344 (69.0%) | 13218 (71.4%) | 1.23 (1.08, 1.17) |
| Index of Multiple Deprivation | Least Deprived | 13598 (70.4%) | 5972 (72.0%) | 1.08 (1.02, 1.14) |
|  | Less Deprived | 19360 (69.3%) | 6244 (71.3%) | 1.10 (1.04, 1.16) |
|  | Middle Quintile | 13623 (68.9%) | 6647 (71.3%) | 1.12 (1.06, 1.18) |
|  | More Deprived | 13627 (69.4%) | 7012 (72.3%) | 1.15 (1.09, 1.22) |
|  | Most Deprived | 14830 (70.8%) | 7631 (73.1%) | 1.12 (1.06, 1.18) |

**Table 2b.** Mean gestational age at abortion before and after the full implementation of home use of misoprostol. (N=145,529)

|  | Population | Pre-implementation period<br>(January 2018 to May 2019)<br>M (sd) | Post-implementation period<br>(June 2019 to December 2019)<br>M (sd) | Risk difference |
| --- | --- | --- | --- | --- |
| **Total** |  | 59.9 (23.0) | 56.8 (21.4) | -3.12 (-3.37, -2.87) |
| **Abortion method** | EMA | 50.1 (8.4) | 48.7 (8.6) | -1.40 (-1.62, -1.17) |
|  | Late medical / surgical | 82.6 (29.3) | 77.63 (29.0) | -4.95 (-5.31, -4.59) |
| **Age (years)** | <18 | 65.7 (26.9) | 62.7 (25.0) | -3.05 (-4.30, -1.80) |
|  | 18-25 | 60.9 (24.0) | 57.5 (22.0) | -3.40 (-3.79, -3.01) |
|  | 26-35 | 58.7 (21.8) | 55.8 (20.5) | -2.91 (-3.29, -2.53) |
|  | 36-45 | 58.6 (22.0) | 56.0 (20.9) | -2.61 (-3.28, -1.94) |
|  | >45 | 57.2 (22.7) | 55.0 (21.2) | -2.18 (-8.28, 3.92) |
| **Place of birth** | England | 59.8 (23.0) | 56.8 (21.5) | -2.96 (-3.24, -2.67) |
|  | European Union | 61.3 (23.7) | 57.7 (21.6) | -3.59 (-4.47, -2.71) |
|  | Northern Ireland | 58.9 (24.4) | 55.5 (18.2) | -3.42 (-9.44, 2.60) |
|  | Outside of EU | 59.7 (22.9) | 56.0 (20.8) | -3.68 (-4.31, -3.04) |
|  | Scotland | 59.3 (23.1) | 56.8 (24.3) | -2.48 (-5.94, 0.97) |
|  | Wales | 61.3 (23.6) | 58.7 (25.6) | -2.63 (-6.00, 0.73) |
| **Disability** | No or Not Reported | 59.8 (23.0) | 56.7 (21.4) | -3.16 (-3.41, -2.91) |
|  | Yes | 62.6 (24.0) | 60.1 (22.8) | -2.53 (-3.85, -1.22) |
| **First language** | English | 59.8 (23.0) | 56.6 (21.4) | -3.13 (-3.38, -2.88) |
|  | Other than English | 64.0 (24.6) | 60.9 (21.8) | -3.08 (-4.45, -1.70) |
| **Race/Ethnicity** | Asian or Asian British | 58.6 (22.6) | 55.1 (20.2) | -3.48 (-4.31, -2.65) |
|  | Black or Black British | 61.4 (24.4) | 57.1 (21.8) | -4.28 (-5.25, -3.31) |
|  | White | 59.8 (23.0) | 56.8 (21.4) | -2.97 (-3.25, -2.68) |
|  | Mixed | 61.2 (23.3) | 57.2 (21.3) | -3.96 (-5.11, -2.82) |
|  | Other/not stated | 61.2 (23.2) | 59.2 (23.8) | -1.99 (-3.50, -0.47) |
| **Religion** | None | 60.0 (23.1) | 56.9 (21.5) | -3.03 (-3.33, -2.72) |
|  | Christian | 60.2 (23.1) | 56.9 (21.4) | -3.25 (-3.80, -2.71) |
|  | Muslim | 59.0 (22.4) | 55.3 (20.7) | -3.70 (-4.78, -2.63) |
|  | Prefer not to say | 60.4 (23.7) | 57.1 (22.1) | -3.30 (-4.62, -1.98) |
|  | Other | 58.2 (22.1) | 55.5 (20.5) | -2.74 (-3.98, -1.50) |
| **Previous abortions** | No previous abortion | 60.3 (23.6) | 57.1 (21.8) | -3.18 (-3.50, -2.87) |
|  | Previous abortion | 59.3 (22.2) | 56.3 (20.8) | -2.99 (-3.39, -2.60) |
| **Index of Multiple Deprivation** | Least Deprived | 58.1 (21.1) | 55.4 (20.1) | -2.70 (-3.28, -2.70) |
|  | Less Deprived | 59.5 (22.6) | 56.4 (21.0) | -3.12 (-3.69, -3.12) |
|  | Middle Quintile | 60.1 (23.3) | 57.1 (22.1) | -3.07 (-3.63, -3.07) |
|  | More Deprived | 60.4 (23.6) | 57.0 (21.6) | -3.39 (-3.94, -3.39) |
|  | Most Deprived | 61.2 (24.3) | 57.7 (22.0) | -3.50 (-4.03, -2.97) |

The median gestational age dropped from 53 days to 50 days following the approval. The gestational age at EMA decreased from 50.1 to 48.7 days [risk difference -1.40 (−1.62, -1.17)]. We also saw a decrease in mean gestation for patients undergoing late medical and surgical abortions: from 82.6 to 77.63 days [-4.95 (−5.31, -4.59)]. Significant decreases were seen across all but the oldest age group; and across all birth places except Wales, Scotland and Northern Ireland. The decrease was more marked among non-White patients; among those of non-Christian religious affiliation; and among those who had previously had an abortion.

### Interrupted Time Series

Interrupted time series graphs of EMA and gestational age are presented in Figures 1a & 1b. There was no evidence of autocorrelation. The ‘slope change’ coefficients are interpreted as the change in slope of the relationship between the outcome (abortion method and gestational age) and time associated with the approval. After the approval, the rate of change of the mean proportion of EMAs each month accelerated by an additional 0.07% (95% CI: - 0.02%, 0.15%). This modelled change in trend suggests an extra 4.2% of abortions would be EMAs at the end of the study period, compared to if pre-approval trends had continued into December 2019. We found strong evidence that the existing decline in mean gestational age since 2015 at BPAS was accelerated post-approval each month by an additional -0.11 days (95% CI: -0.18, -0.03). This change in trend suggests that by the end of the study period, on average, abortions would be carried out 3.4 days earlier compared to if pre-approval trends had continued.

**Figure 1a.**
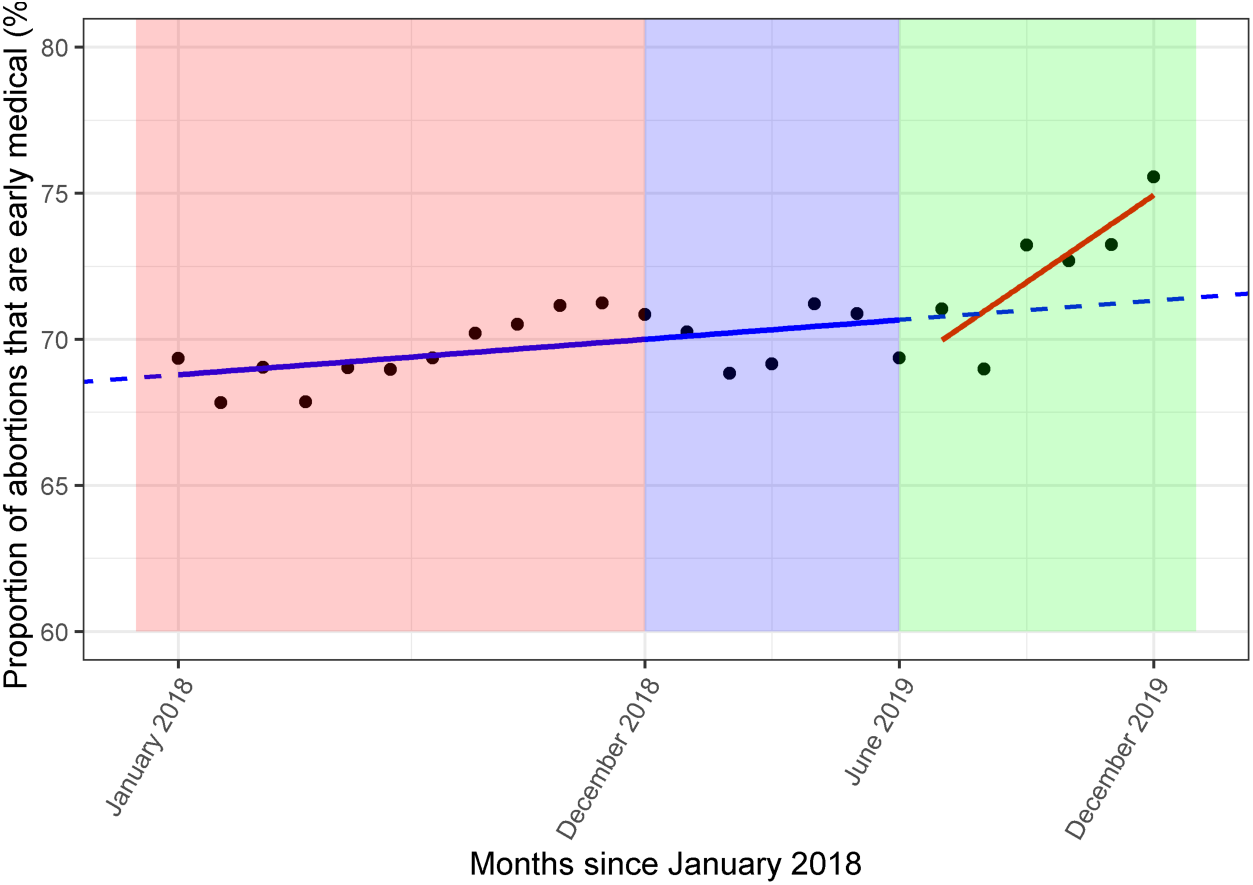
Proportion of all abortions carried out by BPAS that were EMAs, between January 2018 and December 2019 (N=145,529)*

**Figure 1b.**
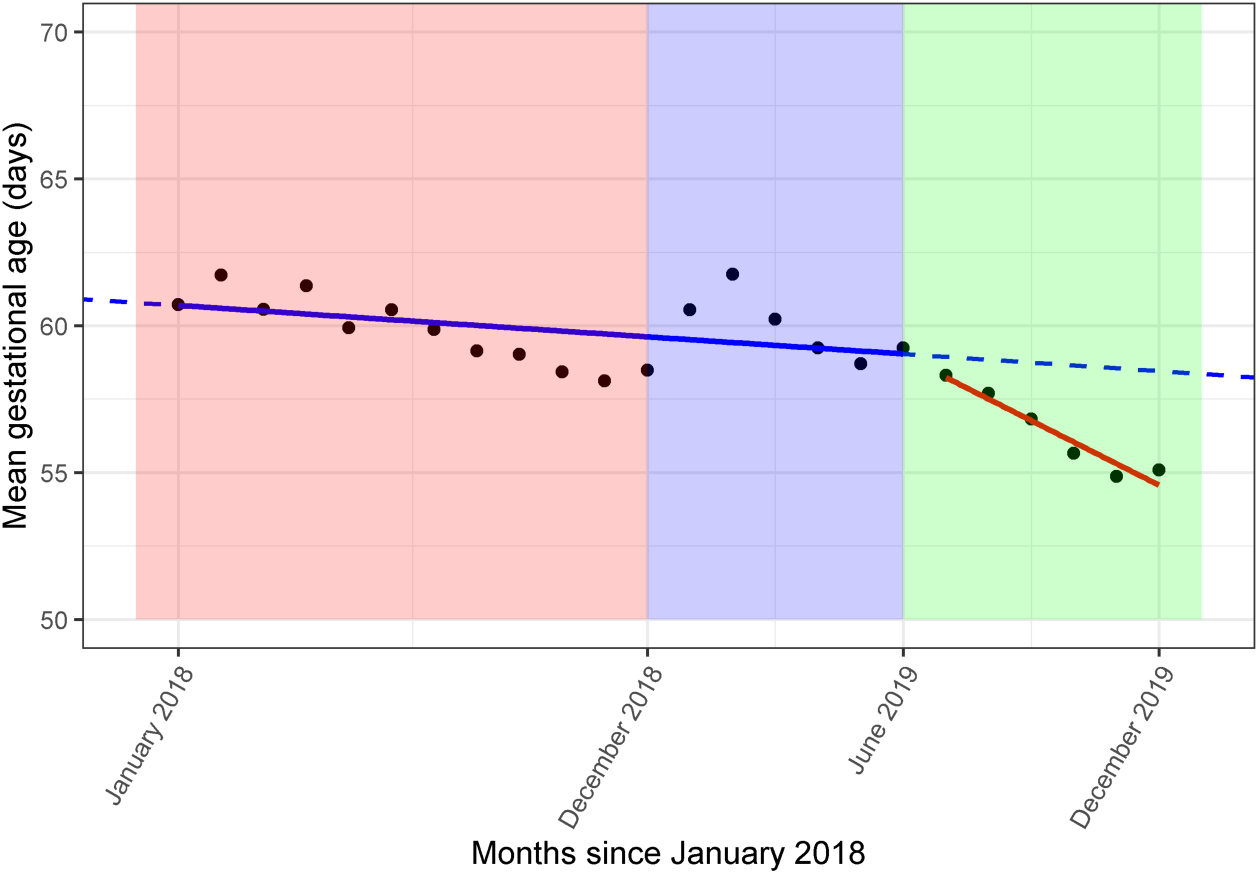
Mean gestational age at abortion, in days, carried out by BPAS between January 2018 and December 2019 (N=145,529)* *The red panel represents time prior to the approval. The approval implementation period is in blue, and the full implementation period is in green. In the time series analyses, the blue implementation period is included with the red pre-approval period.

The estimated slope changes from the stratified time series analyses are presented in Table 3. We found some evidence that the change in slope post-approval differed by levels of race-ethnicity, and by levels of disability for both outcomes, and some weak evidence for difference by IMD. The magnitude of the slope change post-implementation in the proportion of individuals having EMAs and in gestational age generally increased going from least to most deprived. In terms of ethnicity, all groups demonstrated weak evidence of a slope change post-approval on either the EMA measure or the gestational age measure except for Black or Black British women. The largest predicted accelerations in the decrease in gestational age post-approval were seen in Asian or Asian British and White women. Post-approval, the slope change in people with disabilities was faster than in those without disabilities. Figures 2a, 2b, and 2c present illustrative differences in EMAs by IMD, race/ethnicity, and disability.

**Table 3.**
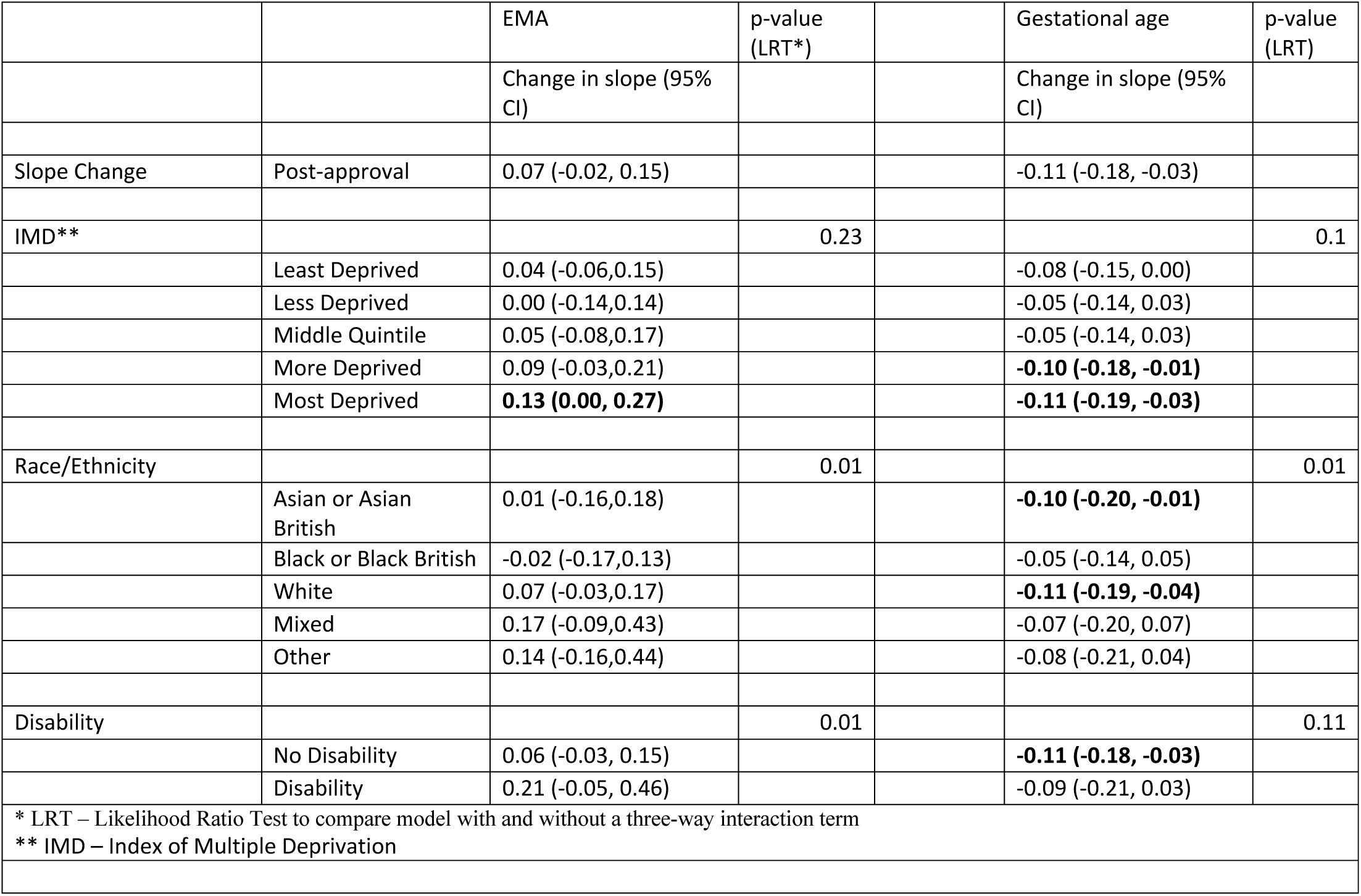
Interrupted Time Series Analyses describing the change in slopes after the implementation (June 2019-December 2019) compared to before (January 2018 – June 2019), of proportion of EMAs and gestational age.

**Figure 2a.**
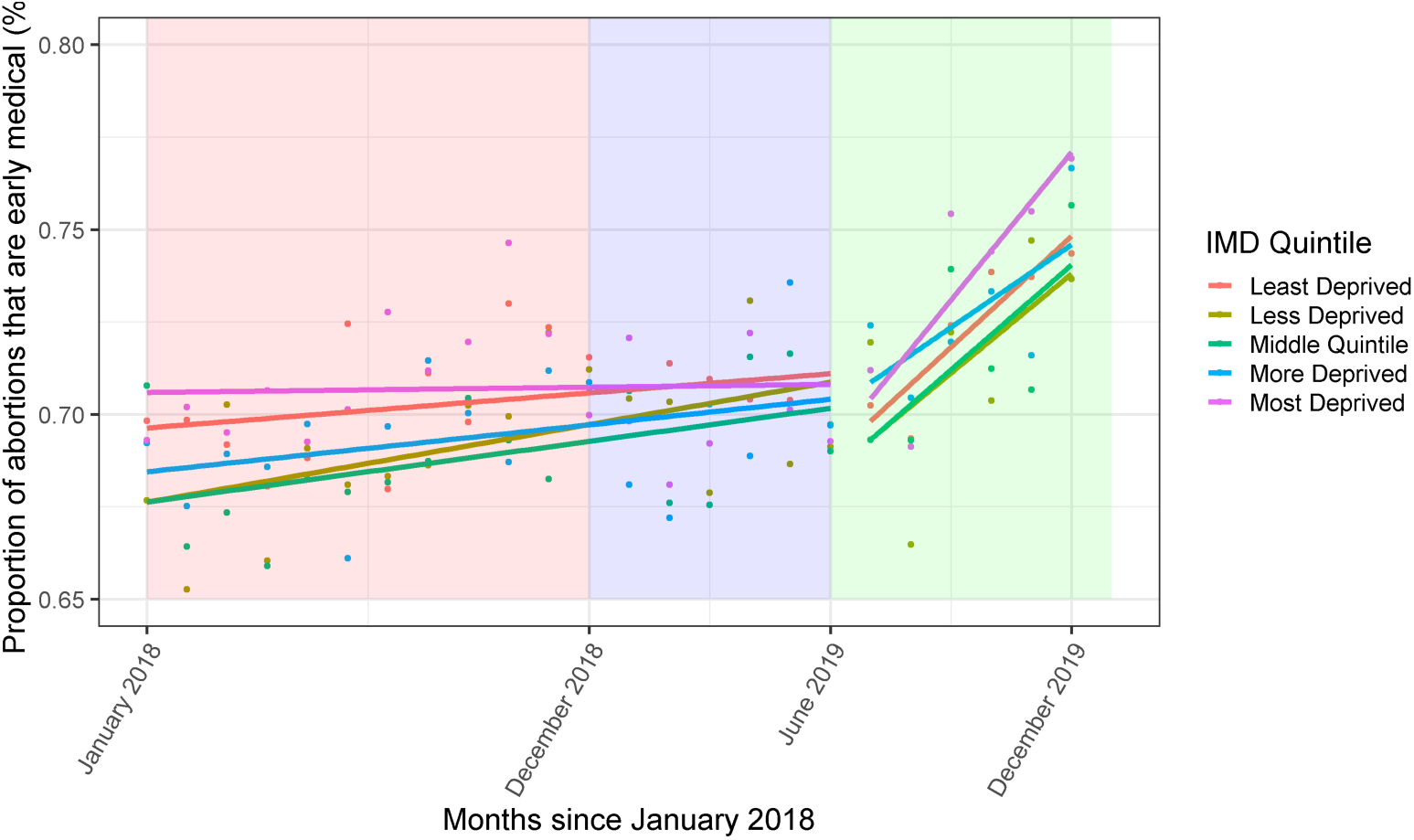
Proportion of all abortions carried out by BPAS that were EMAs, between January 2018 and December 2019, stratified by deprivation quintile. (N=145,529)*

**Figure 2b.**
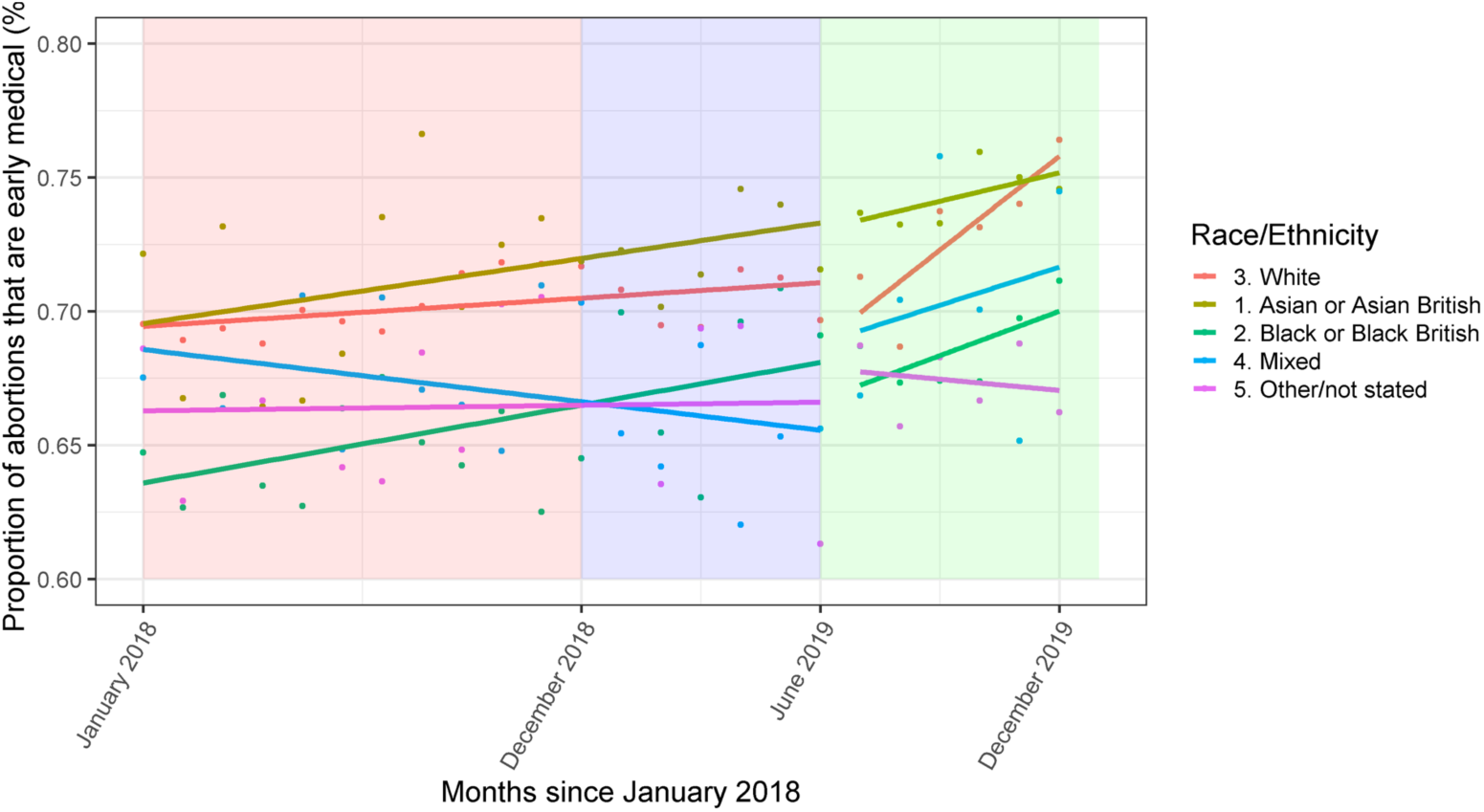
Proportion of all abortions carried out by BPAS that were EMAs, between January 2018 and December 2019, stratified by race/ethnicity. (N=145,529)*

**Figure 2c.**
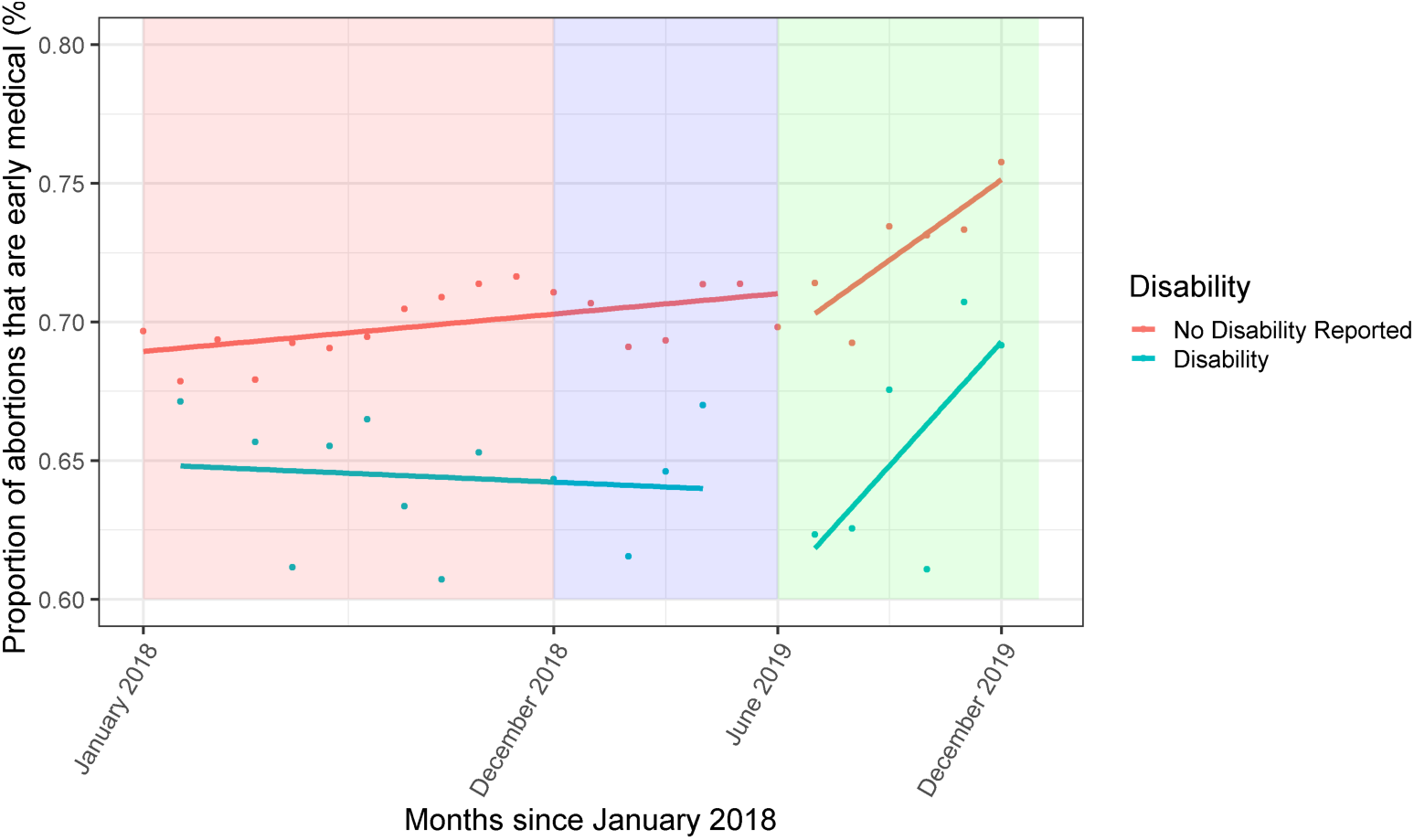
Proportion of all abortions carried out by BPAS that were EMAs, between January 2018 and December 2019, stratified by disability status. (N=145,529)* * The red panel represents time prior to the approval. The approval implementation period is in blue, and the full implementation period is in green. In the time series analyses, the blue implementation period is included with the red pre-approval period.

## Discussion

We found that access to EMA improved after the approval of home misoprostol, as evidenced by the higher proportion of EMAs provided, lower gestational age at treatment, and higher odds of having an EMA across almost all ethnic groups and socioeconomic groups. The only group that did not experience an increase in the odds of having EMA was those born in Northern Ireland.

Already prior to the approval for home use of misoprostol in England as part of an EMA regimen, the proportion of abortions performed medically at BPAS was on the rise and gestational age at treatment was declining, but we found that these trends were accelerated. These accelerations were larger in the most deprived quintiles and in those reporting a disability, but not equal across ethnic groups, with Black and Black British women experiencing little change in trajectories post-approval.

### Strengths and limitations

A strength of the study was that it was a unique opportunity to assess the impact of the 2018 approval of home use of misoprostol prior to the COVID-19 pandemic, when medical abortion became the default method in the first 10 weeks of pregnancy. (25) The study sample was large, constituting 35.67% of all 407,992 abortions carried out in England and Wales between 2018 and 2019. (5,21,26) It was also representative, including a wide range of age groups, ethnicities, religions, and national origins. There is some potential source of selection bias resulting from the fact that we used data from only one clinical provider and so were unable to account for clinic-based differences in the speed of implementation or factors favouring a certain method of abortion.

A limitation of the analysis is that the comparator data collection period following the ruling was short, only six months, which may have resulted in an underestimate of eventual changes. The trends would likely have been stronger had there been more time points to analyse. Any extension of this period beyond March 2020, however, would have been confounded by the changes in abortion regulations contingent on the COVID-19 pandemic that were introduced in April 2020.

### Implications for policy & practice

Our finding that the approval of home use of misoprostol was associated with an acceleration of existing trends in lower gestational age at treatment is important because it means more people in Britain can access EMA at home: it facilitates abortions taking place under the upper gestational age limit imposed by law and it can alleviate the distress caused by longer waiting times to abortion. In addition, the earlier in pregnancy abortions take place, the safer and more effective they are. (1,17) The concomitant acceleration in trend toward more EMAs also has cost implications: EMA is less expensive to provide than surgical abortion, and earlier abortions are associated with a reduction in the need for costly surgical management of complications such as incomplete abortion and continuing pregnancy. (18) NICE estimates that in England a reduction of one day in gestational age at abortion could save £1.6 million per year – our findings that the approval of home use of misoprostol facilitated abortions happening over three days earlier show a significant economic potential of the change. (1,27)

At-home administration of misoprostol can particularly improve access to abortion in more deprived areas, for people with disabilities, and in some ethnic groups, as those populations can suffer from barriers to access, such as issues with travel, taking time off work and arranging childcare, significantly more. Home administration showed little evidence of improving access to abortion in Black and Black British women and further research should be conducted to understand this difference in uptake to better tailor abortion care services to reach marginalised populations.

There are other limitations to the DHSC’s approval. Firstly, the narrow definition of “home” might restrict access to some and doesn’t account for people for whom that location may be undesirable on grounds of safety. (27–29) Secondly, the arbitrary cut-off of 9+6 weeks, which is not reflected in literature, and studies have provided no evidence that home expulsion between 10 and 12 weeks is less effective, less acceptable, or less safe. (1,27,30,31)

Since the approval of home use of misoprostol, the COVID-19 pandemic impelled a sudden and rapid shift towards the telemedical model and the home management of the whole medical abortion process. There is accumulating evidence that, much like with misoprostol, home administration of mifepristone is also safe, effective, and preferred by women. (1,27,30,31) This swift move towards remote care resulted in further improvements in access: data from BPAS from March to June 2020 showed that waiting times for appointments halved, with an average of 4 days, and that average gestation fell by over 7 days, comparing the first half of 2019 to that of 2020. (32) The impact of this change on indices such as area-level deprivation, race/ethnicity, and disability is an area for further exploration.

In addition, it is essential to continue exploring patients’ perspectives of those new models of care. In a qualitative study conducted in England during the COVID-19 pandemic, we found that women were overwhelmingly in favour of home self-management of medical abortion, but some stressed that the option of an in-person interaction with health practitioners should remain available. (33) Another study indicated that some women expressed concerns about whether the symptoms were experiencing during home termination were “normal”. (29)

Greater autonomy in the management of EMA by women themselves has demonstrable clinical and experiential benefits and appears to have the potential to improve access by those who often face the greatest barriers to abortion care. The measures allowing for the home administration of mifepristone were approved on a temporary basis, and are currently set to end in August 2022, despite the abundance of evidence showing it to be a more accessible and acceptable model. Our research shows that remote care has a great potential in improving access and equity of abortion care in the country.

## Data Availability

Data reported in this analysis is not available as it is subject to an ethical permission restricted to the authors of the manuscript

## Acknowledgments

We would like to thank BPAS for providing us with their clinical data.

